# Impact of COVID-19 pandemic on the etiology and characteristics of community-acquired pneumonia among children requiring bronchoalveolar lavage in northern China

**DOI:** 10.1101/2023.03.02.23286686

**Authors:** Rui-han Liu, Yu-yan Zhang, Zhou-hua Lu, Chang-qing Shen, Jin Wang, Qing Zhao, Tong-shu Hou, Feng-hai Niu, Qing-xia Kong, Jun Ning, Lei Yang

## Abstract

**Background:** To investigate the etiology and clinical characteristics of community-acquired pneumonia (CAP) among children requiring bronchoalveolar lavage (BAL) and analyze the impact of the coronavirus disease 2019 (COVID-19) pandemic on the pathogen spectrum and clinical manifestations.

**Methods:** Children <14 years old hospitalized with CAP requiring BLA were enrolled between February 2019 to January 2020 and August 2021 to July 2022. Multiplex reverse transcription polymerase chain reaction (mRT-PCR) was used for pathogen detection. The demographic and clinical characteristics were compared between different pathogen-type infection groups, and before and during the COVID-19 pandemic.

**Results:** Pathogen was detected in 91.66% (1363/1487) children. *Mycoplasma pneumoniae*, adenovirus and human rhinovirus were the most frequently detected pathogens. The frequency of detection of virus infections and co-infections was decreased during the pandemic, but the detection of atypical bacterial infections was increased. The clinical manifestations and the results of CT scans and fiberoptic bronchoscopy showed a significant difference between different types of pathogen infection, and lung inflammation was reduced during the COVID-19 pandemic compared with before the pandemic.

**Conclusions:** *M. pneumoniae* infection might be the greatest pediatric disease burden leading to CAP in northern China. Wearing masks and social distancing in public places during the COVID-19 pandemic effectively reduced the transmission of respiratory viruses, but it did not reduce the infection rate of *M. pneumoniae*. In addition, these interventions significantly reduced lung inflammation in children compared with before the pandemic.

## Introduction

Community-acquired pneumonia (CAP) is a common infection that presents a serious public health threat among children under 5 years of age. CAP was responsible for 13.9% of childhood deaths among the under-5s worldwide in 2019[1]. Although this was more than 1,001,000 fewer deaths compared with the year 2000, the number of complicated and severe pneumonia cases has increased considerably over the past two decades[2,3]. Following the introduction of the pneumococcal conjugate vaccine and the *Haemophilus influenzae* type b vaccine, respiratory viruses and atypical bacteria have become the most frequent etiologic agents in recent years[4].

The outbreak of coronavirus disease 2019 (COVID-19) constitutes a substantial health burden across the globe. To control the COVID-19 pandemic, governments around the world implemented a series of non-pharmaceutical interventions (NPIs), such as mandated facial masks, encouraged social distancing, remote working, online teaching, stay-at-home orders and travel restrictions. Studies have shown that the preventive measures useful in containing COVID-19 also seemed to be effective in preventing the transmission of other respiratory viruses[5-11]. Considering the impact of etiology on disease severity, mortality and long-term sequelae in children with CAP[12], it is necessary to identify the causative pathogen.

Nasopharyngeal swabs or aspirate specimens are usually used for pathogen detection in children with CAP. However, the detection of microorganisms from the upper respiratory tract may not directly correlate with the pathogenic agents of pneumonia[13]. Over the past two decades, fiberoptic bronchoscopy (FB) and bronchoalveolar lavage (BAL) have become important diagnostic and therapeutic procedures for severe pneumonia. Bronchoalveolar lavage fluid (BALF) collected directly from the foci of inflammation can be used as reliable samples for pathogen identification[14]. Furthermore, advances in molecular detection technology have contributed to the rapid and accurate pathogen identification. Automated capillary electrophoresis combined with multiplex reverse transcription polymerase chain reaction (mRT-PCR) has been developed to detect microorganisms, at reasonable cost and acceptable levels of sensitivity and specificity[15].

In this study, we report and discuss the impact of the COVID-19 pandemic on the spectrum of common respiratory pathogens among children with CAP requiring FB plus BAL in northern China. Additionally, changes in the pathogen spectrum and the clinical manifestations in the children were analyzed systematically and compared prior to the COVID-19 pandemic (February 2019 to January 2020) and during the COVID-19 pandemic (August 2021 to July 2022).

## Materials and methods

### Ethics approval

This study was approved by the Ethics Committee of the Affiliated Hospital of Jining Medical University (No.: 2020C076).

### Study population

Between February 2019 to January 2020 and August 2021 to July 2022, a total of 1487 children under the age of 14 who required BAL were included in our study. It was ensured by experienced physicians that the following inclusion criteria were met. (1) All of the children were in the acute stage of pneumonia. (2) Symptoms persisted for at least 7 days after the initiation of treatment and/or the patient was not responding to treatment, or breathing was labored, or the child became distressed or agitated, or their oxygen saturation level was < 92%. (3) Auscultation revealed decreased/absent breath sounds, or tubular breath sounds, and or a repeated fixed wheeze. (4) Radiographic inspection revealed pulmonary atelectasis, or pulmonary emphysema, or mediastinal emphysema, or pulmonary consolidation. Exclusion criteria were as follows: children with immunodeficiency disease, congenital heart disease, tracheostomy tube, cystic fibrosis, hemopoietic system malignant tumor, recent hematopoietic stem-cell or solid-organ transplant, children with an alternative respiratory disorder diagnosis or newborns less than 1 month old. Written informed consent was obtained from parents/guardians before children underwent FB plus BAL.

### Sample and data collection

For the purpose of treatment, FB combined with BAL was performed by experienced bronchoscopists according to the recommendation of the European Respiratory Society Task Force[16]. Approximately 3 mL of BALF was collected from each patient and the samples were aliquoted and stored at −80°C within 2 hours simultaneously. Diagnosis of the morphology under bronchoscopy was made by two experienced physicians[17,18].

The presence of pulmonary consolidation, infiltration, pleural effusion and pericardial effusion in the CT scans was considered evidence of pneumonia[19]. The CT scans were reviewed by three senior radiologists independently to ensure the accuracy and consistency of the diagnosis. The other clinical data were extracted from electronic medical records after the patient was discharged.

### Pathogen detection

The SureX 13 respiratory pathogen multiplex kit (Ningbo Health Gene Technologies Ltd.) was used to simultaneously detect common virus and atypical bacteria in 200 µL of BALF. The mRT-PCR products were analyzed on the Applied Biosystems 3500Dx Genetic Analyzer with LIZ500 as the internal standard, and then analyzed with GeneMapper 4.1 software (Thermo Fisher Scientific, Waltham, MA, USA).

A viral or atypical bacterial pathogen was considered to be present if influenza A virus (Flu A, including 2009 H1N1 pandemic and H3N2), influenza B virus (Flu B), parainfluenza virus (PIV), adenovirus (AdV), respiratory syncytial virus (RSV), human rhinovirus (HRV), human metapneumovirus (hMPV), human bocavirus (HBoV), coronavirus (CoV), *Mycoplasma pneumoniae* or *Chlamydophila pneumoniae* was detected in the BALF.

Co-infection was considered when two or more viral or atypical bacterial pathogens were detected in one child.

### Statistical analysis

Considering the similar clinical features observed between children with virus–virus co-infections and single virus infections, these cases were combined into the virus group in the tables. This also applied to those with one or two atypical bacterial infections, who were combined in the atypical bacterium group. The clinical characteristics of enrolled children were stratified and described according to the following groups: virus or atypical bacterium only infection, and co-infection (virus–atypical bacterium infections).

All statistical analyses were performed using SAS software, version 9.4 (SAS Institute Inc., NC, USA). The median and quartile [M (P25, P75)] values were used to describe quantitative data with a non-normal distribution, and the frequency (component ratio) [n (%)] was used to describe classified data. The Chi-squared test was used to analyze differences in sex, age, the season of disease onset and the infecting pathogens between samples collected before and during the COVID-19 pandemic. The Kruskal–Wallis H test was used to compare the time from illness onset to admission, the duration of hospitalization, and the presence of a fever and cough in different groups between different time periods. Fisher’s exact test was used to compare the difference in the proportion of different pathogens between samples taken before and during the COVID-19 pandemic. Trends in the infection rates of various pathogens in different age groups were assessed by the Cochran–Armitage trend test. The differences in clinical manifestations, CT scans and FB results between the various groups and different time periods were also analyzed by Fisher’s exact test. A probability (p) value less than 0.05 was considered statistically significant. For pairwise comparison of multiple groups of samples, Bonferroni correction was used to inspect the standard of each comparison, α′= 0.01667 (0.05/3).

## Results

### Demographic characteristics

From February 2019 to January 2020 and August 2021 to July 2022, 696 and 791 children were enrolled in this study, respectively (Table 1). A total of 54.74% of the children were boys, and the median age was 70.51±32.79 months. The median time from illness onset to admission and the median duration of hospitalization were 8 (6,10) days and 7 (6,9) days, respectively. The proportion of children undergoing FB combined with BAL gradually increased with age (X^2^/Z=54.0288, P<0.001), and the proportion of children affected during autumn was significantly higher than that in the other seasons.

**Table 1.**
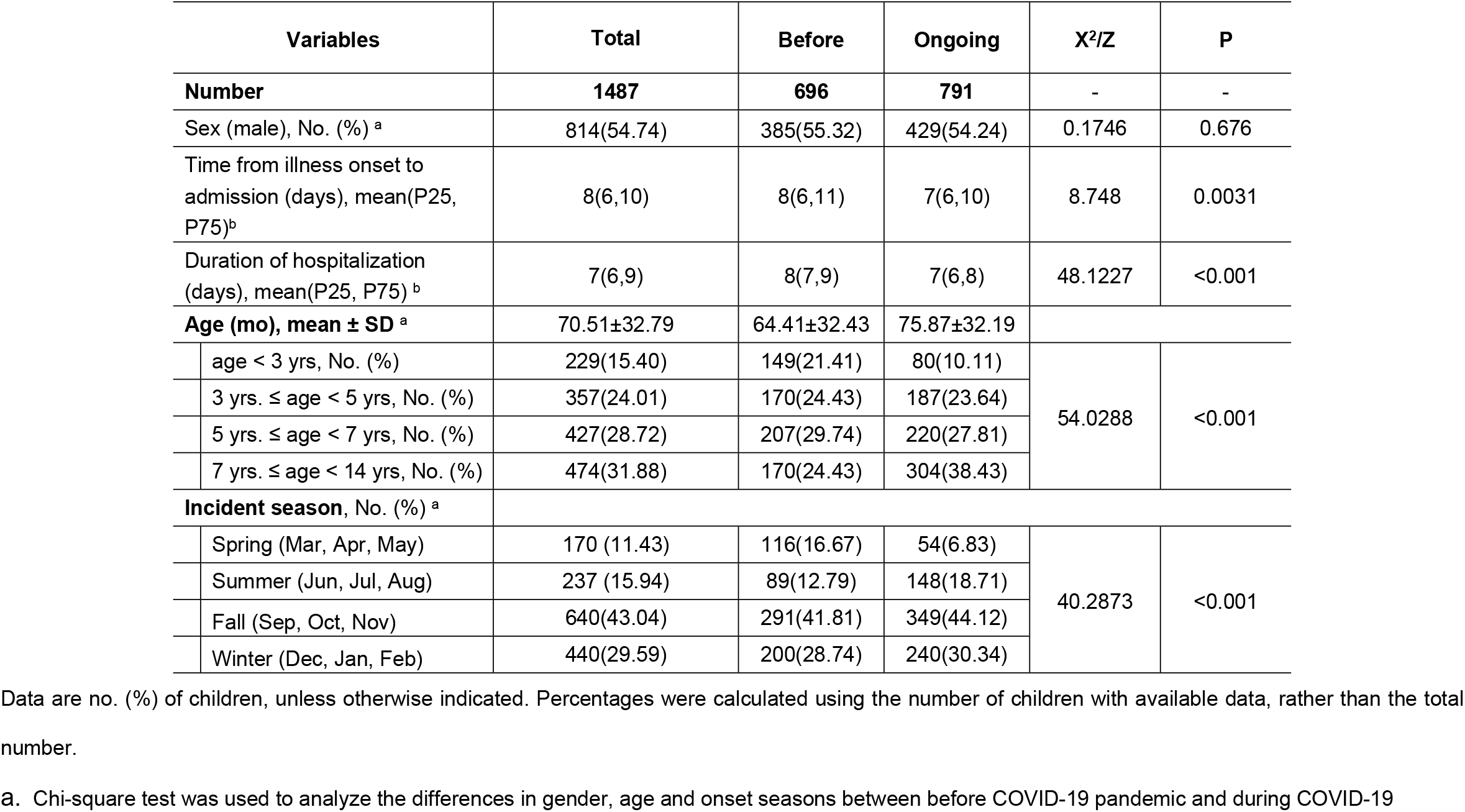
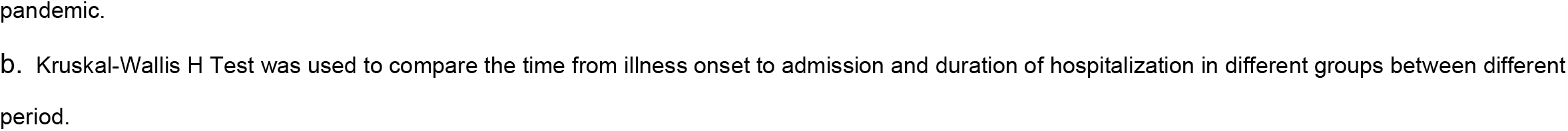
Demographic characteristics of children with community-acquired pneumonia requiring Flexible bronchoscopy and bronchoalveolar lavage.

| Variables |  | Total | Before | Ongoing | X<sup>2</sup>/Z | P |
| --- | --- | --- | --- | --- | --- | --- |
| Number |  | 1487 | 696 | 791 | - | - |
| Sex (male), No. (%) <sup>a</sup> |  | 814(54.74) | 385(55.32) | 429(54.24) | 0.1746 | 0.676 |
| Time from illness onset to admission (days), mean(P25, P75)<sup>b</sup> |  | 8(6,10) | 8(6,11) | 7(6,10) | 8.748 | 0.0031 |
| Duration of hospitalization (days), mean(P25, P75) <sup>b</sup> |  | 7(6,9) | 8(7,9) | 7(6,8) | 48.1227 | <0.001 |
| Age (mo), mean ± SD <sup>a</sup> |  | 70.51±32.79 | 64.41±32.43 | 75.87±32.19 |  |  |
|  | age < 3 yrs, No. (%) | 229(15.40) | 149(21.41) | 80(10.11) | 54.0288 | <0.001 |
|  | 3 yrs. ≤ age < 5 yrs, No. (%) | 357(24.01) | 170(24.43) | 187(23.64) |  |  |
|  | 5 yrs. ≤ age < 7 yrs, No. (%) | 427(28.72) | 207(29.74) | 220(27.81) |  |  |
|  | 7 yrs. ≤ age < 14 yrs, No. (%) | 474(31.88) | 170(24.43) | 304(38.43) |  |  |
| Incident season, No. (%) <sup>a</sup> |  |  |  |  |  |  |
|  | Spring (Mar, Apr, May) | 170 (11.43) | 116(16.67) | 54(6.83) | 40.2873 | <0.001 |
|  | Summer (Jun, Jul, Aug) | 237 (15.94) | 89(12.79) | 148(18.71) |  |  |
|  | Fall (Sep, Oct, Nov) | 640(43.04) | 291(41.81) | 349(44.12) |  |  |
|  | Winter (Dec, Jan, Feb) | 440(29.59) | 200(28.74) | 240(30.34) |  |  |

Data are no. (%) of children, unless otherwise indicated. Percentages were calculated using the number of children with available data, rather than the total number.
**a.** Chi-square test was used to analyze the differences in gender, age and onset seasons between before COVID-19 pandemic and during COVID-19
pandemic.
**b.** Kruskal-Wallis H Test was used to compare the time from illness onset to admission and duration of hospitalization in different groups between different period.

Compared with before the COVID-19 pandemic, the median time from illness onset to hospitalization was reduced [8 (6,11) days vs 7 (6,10) days; P=0.0031] and the median hospitalization time was also reduced [8 (7,9) days vs 7 (6,8) days; P<0.001] in the children enrolled during the COVID-19 pandemic. During the COVID-19 pandemic, the pathogen detection rate decreased from 20% to ∼10% in children under 3 years of age and increased from 25% up to ∼40% in children aged over 7 years. There was also a statistical difference in the season of illness onset among the children (X²/Z=40.2873, P<0.001), with a lower incidence of disease in spring (16.67% vs 6.83%) and a higher incidence in summer (12.79% vs 18.71%) during the COVID-19 pandemic (Table 1).

### Pathogen spectrum

Among the enrolled children, ≥1 pathogen was detected in 91.66% (1363/1487) of children and the pathogen detection rate gradually increasing with age (Z=−6.425, P<0.001). The specific pathogens detected are listed in Table 2, sorted by descending detection rates. AdV (n=64, 4.30%), HRV (n=41, 2.76%), PIV (n=21, 1.41%) and Flu B (n=21, 1.41%) were the most common viral pathogens among the enrolled cases. There were statistically significant differences in the age of children infected with viruses, with the number of children infected with viruses decreasing with increased age, except for Flu B (P=0.1927) and hMPV (P=0.1447). Children younger than 3 years were more vulnerable to AdV, PIV and RSV infection, whereas children older than 3 years were more susceptible to atypical bacterial infection. A total of 1254 (84.33%) cases were found to be infected with *M. pneumoniae*, and the number of children with *M. pneumoniae* infection increased with age (Z=−10.2742, P<0.001).

**Table 2.** Prevalence of causative respiratory pathogen according to age groups in children performed with FOB combined with BAL.

| **Pathogen Detected** | **Total**<br>**No. (%)** | **< 3 years**<br>**No. (%)** | **3–5 years**<br>**No. (%)** | **5–7 years**<br>**No. (%)** | **7–14 years**<br>**No. (%)** | **Z** | **p** |
| --- | --- | --- | --- | --- | --- | --- | --- |
| **Participants, n** | **1487** | **229** | **357** | **427** | **474** | **-** | **-** |
| ≥1 pathogen detected | 1363(91.66) | 186(81.22) | 319(89.36) | 407(95.32) | 451(95.15) | -6.425 | <0.001 |
| **Virus** |  |  |  |  |  |  |  |
| AdV <sup>a</sup> | 64(4.30) | 19(8.30) | 25(7.00) | 13(3.04) | 7(1.48) | 4.9844 | <0.001 |
| HRV <sup>a</sup> | 41(2.76) | 10(4.37) | 11(3.08) | 11(2.58) | 9(1.90) | 1.8833 | 0.0298 |
| PIV <sup>a</sup> | 21(1.41) | 12(5.24) | 5(1.40) | 2(0.47) | 2(0.42) | 4.6023 | <0.001 |
| Flu B | 21(1.41) | 5(2.18) | 2(0.56) | 11(2.58) | 3(0.63) | 0.8681 | 0.1927 |
| RSV <sup>a</sup> | 19(1.28) | 11(4.80) | 2(0.56) | 4(0.94) | 2(0.42) | 3.8454 | <0.001 |
| Flu A <sup>a</sup> | 10(0.67) | 6(2.62) | 2(0.56) | 0(0.00) | 2(0.42) | 2.9073 | 0.0018 |
| HBoV <sup>a</sup> | 8(0.54) | 5(2.18) | 1(0.28) | 1(0.23) | 1(0.21) | 2.7325 | 0.0031 |
| hMPV | 8(0.54) | 2(0.87) | 2(0.56) | 3(0.7) | 1(0.21) | 1.0593 | 0.1447 |
| CoV <sup>a</sup> | 2(0.13) | 1(0.44) | 1(0.28) | 0(0.00) | 0(0.00) | 1.6974 | 0.0448 |
| **Atypical bacteria** |  |  |  |  |  |  |  |
| *M.pneumonia* <sup>a</sup> | 1254(84.33) | 142(62.01) | 291(81.51) | 381(89.23) | 440(92.83) | -10.2742 | <0.001 |
| *C.pneumoniae* | 4(0.27) | 1(0.44) | 0(0.00) | 1(0.23) | 2(0.42) | -0.4335 | 0.3323 |

Data are no. (%) of children, unless otherwise indicated. Percentages were calculated using the number of children with available data, rather than the total number.
<sup>a</sup> $p < 0.05$ , Cochran-Armitage Trend Test was used to compare trends of infection rates of various pathogens in different age groups.

Compared with before the COVID-19 pandemic, the number of children infected with AdV (5.75% vs 3.03%, P=0.0105), HRV (4.17% vs 1.52%, P=0.0022), Flu B (2.16% vs 0.76%, P=0.0271), Flu A (1.44% vs 0.00%, P=0.0005) and HBoV (1.01% vs 0.13%, P=0.0293) decreased significantly. Therefore, the total proportion of cases of viral infection (9.91% vs 4.80%) and co-infection (7.33% vs 3.54%) decreased significantly. Whereas the proportion of cases of *M. pneumoniae* infection had a further increasing that compared with before the pandemic (87.23% vs 81.03%, P=0.0013) (Table 3).

**Table 3.**
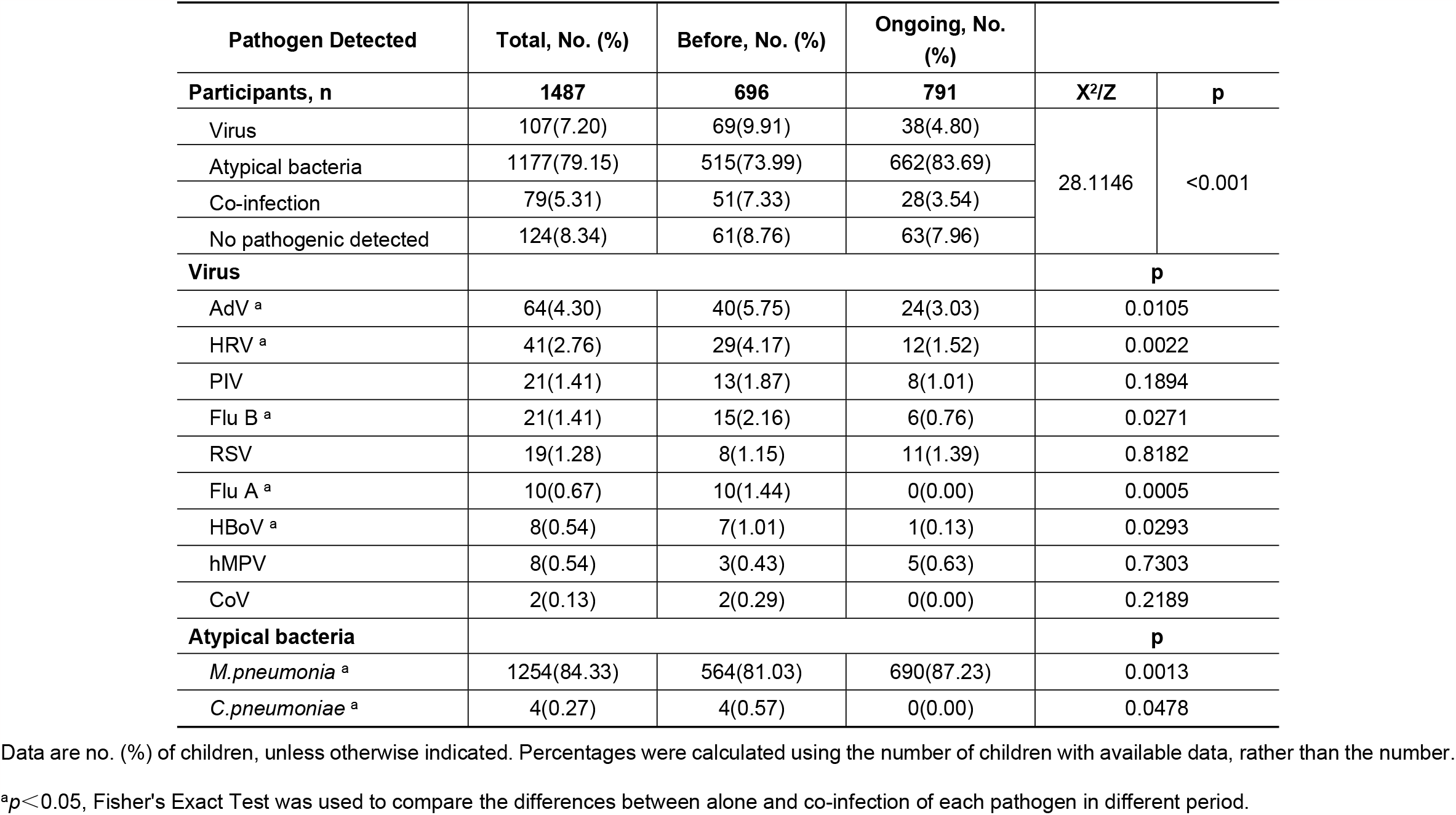
Distribution of various pathogens in children patients performed with FOB combined with BAL.

### Clinical characteristics between three infection groups

Children with viral infection showed the shortest duration of fever [4 (0,6) days], while those with atypical bacterial infection [7 (5,9) days vs 4 (0,6) days, P<0.001] or co-infection [7 (4,10) days vs 4 (0,6) days, P=0.0002] showed longer duration of fever, and the differences among the three groups were statistically significant (P<0.001). The incidence of wheezing in children infected with virus (18.69% vs 3.91%, P<0.001) and those with co-infection (15.19% vs 3.91%, P=0.0001) was significantly higher than in those with atypical bacterial infection (3.91%). Compared with the atypical bacterial infection group, viral infection made children more prone to dyspnea (8.41% vs 1.44%, P=0.0001), diarrhea (15.89% vs 6.73%, P=0.0019), rhonchi (47.66% vs 28.63%, P=0.0003) and chest indrawing (9.35% vs 1.7%, P=0.0001). Decreased breath sounds (59.64%) and rales (66.10%) were the most common signs in the atypical bacterial infection group. No remarkable differences in the duration of cough, fatigue, lack of appetite, abdominal pain, chest pain and tachypnea were observed between the three groups (Table 4). In CT scan findings, both consolidation and infiltration revealed prominent differences among the different infection groups. Atypical bacterial infection was most likely to cause consolidation (P<0.001) and single lobar infiltration (P=0.0222). Compared with the atypical bacterial infection group, the virus infection group was more likely to lead to bilateral multilobar infiltration (46.73% vs 30.42%, P=0.0007), whereas unilateral multilobar infiltration was more common in the atypical bacterial infection group than in the virus infection group (28.29% vs 17.76%, P=0.0177) or the co-infection group (31.65% vs 17.76%, P=0.0359) (Table 4).

**Table 4.**
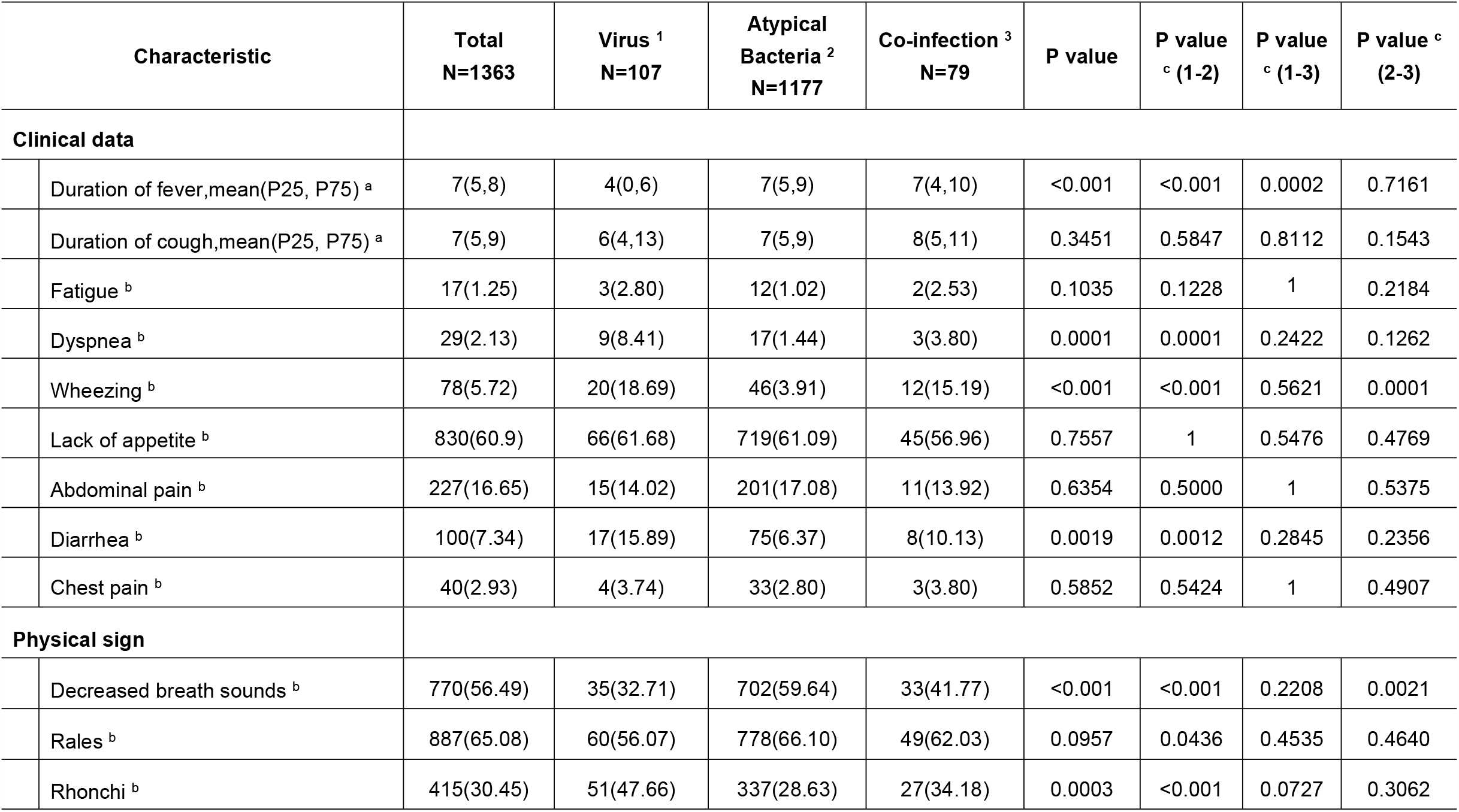
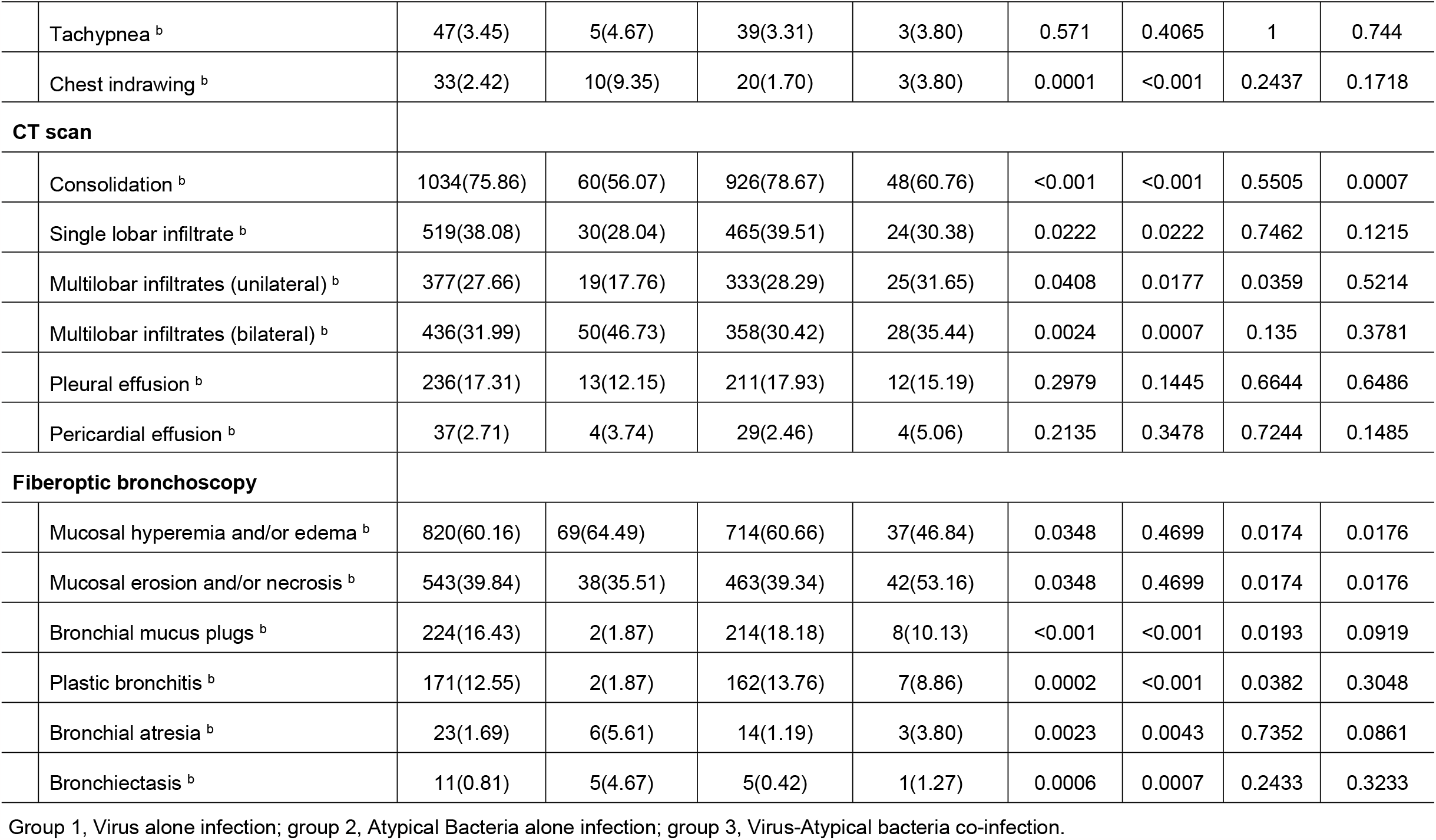
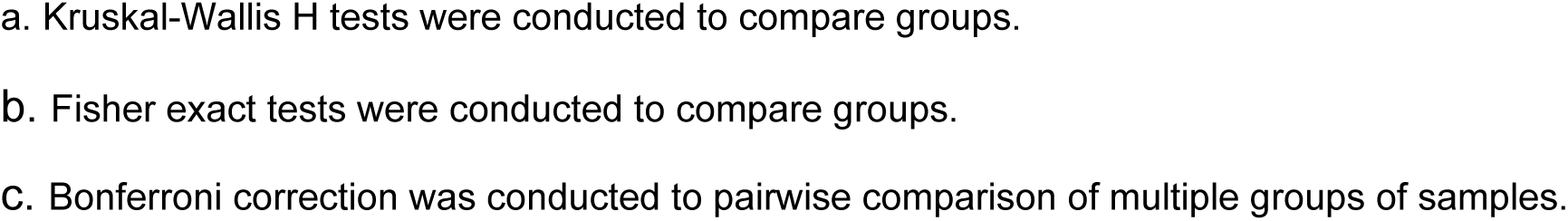
Comparison of clinical and laboratory findings according to respiratory pathogens in children performed with FOB combined with BAL.

| Characteristic |  | Total<br>N=1363 | Virus <sup>1</sup><br>N=107 | Atypical<br>Bacteria <sup>2</sup><br>N=1177 | Co-infection <sup>3</sup><br>N=79 | P value | P value<br><sup>c</sup> (1-2) | P value<br><sup>c</sup> (1-3) | P value <sup>c</sup><br>(2-3) |
| --- | --- | --- | --- | --- | --- | --- | --- | --- | --- |
| **Clinical data** |  |  |  |  |  |  |  |  |  |
|  | Duration of fever,mean(P25, P75) <sup>a</sup> | 7(5,8) | 4(0,6) | 7(5,9) | 7(4,10) | <0.001 | <0.001 | 0.0002 | 0.7161 |
|  | Duration of cough,mean(P25, P75) <sup>a</sup> | 7(5,9) | 6(4,13) | 7(5,9) | 8(5,11) | 0.3451 | 0.5847 | 0.8112 | 0.1543 |
|  | Fatigue <sup>b</sup> | 17(1.25) | 3(2.80) | 12(1.02) | 2(2.53) | 0.1035 | 0.1228 | 1 | 0.2184 |
|  | Dyspnea <sup>b</sup> | 29(2.13) | 9(8.41) | 17(1.44) | 3(3.80) | 0.0001 | 0.0001 | 0.2422 | 0.1262 |
|  | Wheezing <sup>b</sup> | 78(5.72) | 20(18.69) | 46(3.91) | 12(15.19) | <0.001 | <0.001 | 0.5621 | 0.0001 |
|  | Lack of appetite <sup>b</sup> | 830(60.9) | 66(61.68) | 719(61.09) | 45(56.96) | 0.7557 | 1 | 0.5476 | 0.4769 |
|  | Abdominal pain <sup>b</sup> | 227(16.65) | 15(14.02) | 201(17.08) | 11(13.92) | 0.6354 | 0.5000 | 1 | 0.5375 |
|  | Diarrhea <sup>b</sup> | 100(7.34) | 17(15.89) | 75(6.37) | 8(10.13) | 0.0019 | 0.0012 | 0.2845 | 0.2356 |
|  | Chest pain <sup>b</sup> | 40(2.93) | 4(3.74) | 33(2.80) | 3(3.80) | 0.5852 | 0.5424 | 1 | 0.4907 |
| **Physical sign** |  |  |  |  |  |  |  |  |  |
|  | Decreased breath sounds <sup>b</sup> | 770(56.49) | 35(32.71) | 702(59.64) | 33(41.77) | <0.001 | <0.001 | 0.2208 | 0.0021 |
|  | Rales <sup>b</sup> | 887(65.08) | 60(56.07) | 778(66.10) | 49(62.03) | 0.0957 | 0.0436 | 0.4535 | 0.4640 |
|  | Rhonchi <sup>b</sup> | 415(30.45) | 51(47.66) | 337(28.63) | 27(34.18) | 0.0003 | <0.001 | 0.0727 | 0.3062 |
|  | Tachypnea <sup>b</sup> | 47(3.45) | 5(4.67) | 39(3.31) | 3(3.80) | 0.571 | 0.4065 | 1 | 0.744 |
|  | Chest indrawing <sup>b</sup> | 33(2.42) | 10(9.35) | 20(1.70) | 3(3.80) | 0.0001 | <0.001 | 0.2437 | 0.1718 |
| **CT scan** |  |  |  |  |  |  |  |  |  |
|  | Consolidation <sup>b</sup> | 1034(75.86) | 60(56.07) | 926(78.67) | 48(60.76) | <0.001 | <0.001 | 0.5505 | 0.0007 |
|  | Single lobar infiltrate <sup>b</sup> | 519(38.08) | 30(28.04) | 465(39.51) | 24(30.38) | 0.0222 | 0.0222 | 0.7462 | 0.1215 |
|  | Multilobar infiltrates (unilateral) <sup>b</sup> | 377(27.66) | 19(17.76) | 333(28.29) | 25(31.65) | 0.0408 | 0.0177 | 0.0359 | 0.5214 |
|  | Multilobar infiltrates (bilateral) <sup>b</sup> | 436(31.99) | 50(46.73) | 358(30.42) | 28(35.44) | 0.0024 | 0.0007 | 0.135 | 0.3781 |
|  | Pleural effusion <sup>b</sup> | 236(17.31) | 13(12.15) | 211(17.93) | 12(15.19) | 0.2979 | 0.1445 | 0.6644 | 0.6486 |
|  | Pericardial effusion <sup>b</sup> | 37(2.71) | 4(3.74) | 29(2.46) | 4(5.06) | 0.2135 | 0.3478 | 0.7244 | 0.1485 |
| **Fiberoptic bronchoscopy** |  |  |  |  |  |  |  |  |  |
|  | Mucosal hyperemia and/or edema <sup>b</sup> | 820(60.16) | 69(64.49) | 714(60.66) | 37(46.84) | 0.0348 | 0.4699 | 0.0174 | 0.0176 |
|  | Mucosal erosion and/or necrosis <sup>b</sup> | 543(39.84) | 38(35.51) | 463(39.34) | 42(53.16) | 0.0348 | 0.4699 | 0.0174 | 0.0176 |
|  | Bronchial mucus plugs <sup>b</sup> | 224(16.43) | 2(1.87) | 214(18.18) | 8(10.13) | <0.001 | <0.001 | 0.0193 | 0.0919 |
|  | Plastic bronchitis <sup>b</sup> | 171(12.55) | 2(1.87) | 162(13.76) | 7(8.86) | 0.0002 | <0.001 | 0.0382 | 0.3048 |
|  | Bronchial atresia <sup>b</sup> | 23(1.69) | 6(5.61) | 14(1.19) | 3(3.80) | 0.0023 | 0.0043 | 0.7352 | 0.0861 |
|  | Bronchiectasis <sup>b</sup> | 11(0.81) | 5(4.67) | 5(0.42) | 1(1.27) | 0.0006 | 0.0007 | 0.2433 | 0.3233 |

Group 1, Virus alone infection; group 2, Atypical Bacteria alone infection; group 3, Virus-Atypical bacteria co-infection.
**a.** Kruskal-Wallis H tests were conducted to compare groups. **b.** Fisher exact tests were conducted to compare groups. **c.** Bonferroni correction was conducted to pairwise comparison of multiple groups of samples.

The FB findings showed that compared with the viral infection and atypical bacterial infection groups, the co-infection group presented more severe tracheal mucosal injury. The proportion of more severe mucosal erosion and/or necrosis was the highest in co-infection group (53.16% vs 35.51%, P=0.0174; 53.16% vs 39.34%, P=0.0176). Compared with the viral infection group, bronchial mucus plugs and plastic bronchitis were more frequently detected in children with atypical bacterial infections (18.18% vs 1.87%, P<0.001) and co-infections (10.13% vs 1.87%, P=0.0193). Compared with the atypical bacterial infection group, virus infection was more likely to give rise to alterations in bronchial lumen morphology, which included bronchial atresia (5.61% vs 1.19%, P=0.0043) and bronchiectasis (4.67% vs 0.42%, P=0.0007) (Table 4).

### Clinical characteristics before and during the COVID-19 pandemic

Compared with before the COVID-19 pandemic, clinical symptoms and pulmonary signs in the enrolled children during the pandemic showed a shorter duration of cough [7 (5,10) days vs 6 (4,9) days; P<0.001], a decreased proportion of rhonchi dyspnea (38.11% vs 23.76%, P<0.001) and chest indrawing (3.46% vs 1.51%, P=0.0216), but an increased proportion of chest pain (1.89% vs 3.85%, P=0.0364), decreased breath sounds (47.87% vs 64.01%, P<0.001) and rales (49.92% vs 78.30%, P<0.001). CT scans revealed that the proportions of consolidation (91.76% vs 57.64%, P<0.001) and unilateral multilobar infiltrates (32.55% vs 22.05%, P<0.001) were higher than before the COVID-19 pandemic, but the proportions of bilateral multilobar infiltrates (28.57% vs 35.91%, P=0.0043) and pleural effusion (13.74% vs 21.42%, P=0.0002) were lower than before the COVID-19 pandemic. The FB findings revealed an increased proportion of mucosal hyperemia and/or edema (27.09% vs 89.01%, P<0.001), which corresponded to the decreased proportion of mucosal erosion and/or necrosis (72.91% vs 10.99%, P<0.001). Despite a significant increase in the proportion of bronchial mucus plugs (13.70% vs 18.82%, P=0.0126), there was a significant reduction in bronchial atresia (3.31% vs 0.27%, P<0.001) and bronchiectasis (1.57% vs 0.14%, P=0.0039). The above results suggest that lung inflammation in the enrolled children was significantly alleviated compared with before the COVID-19 pandemic (Table 5).

**Table 5.**
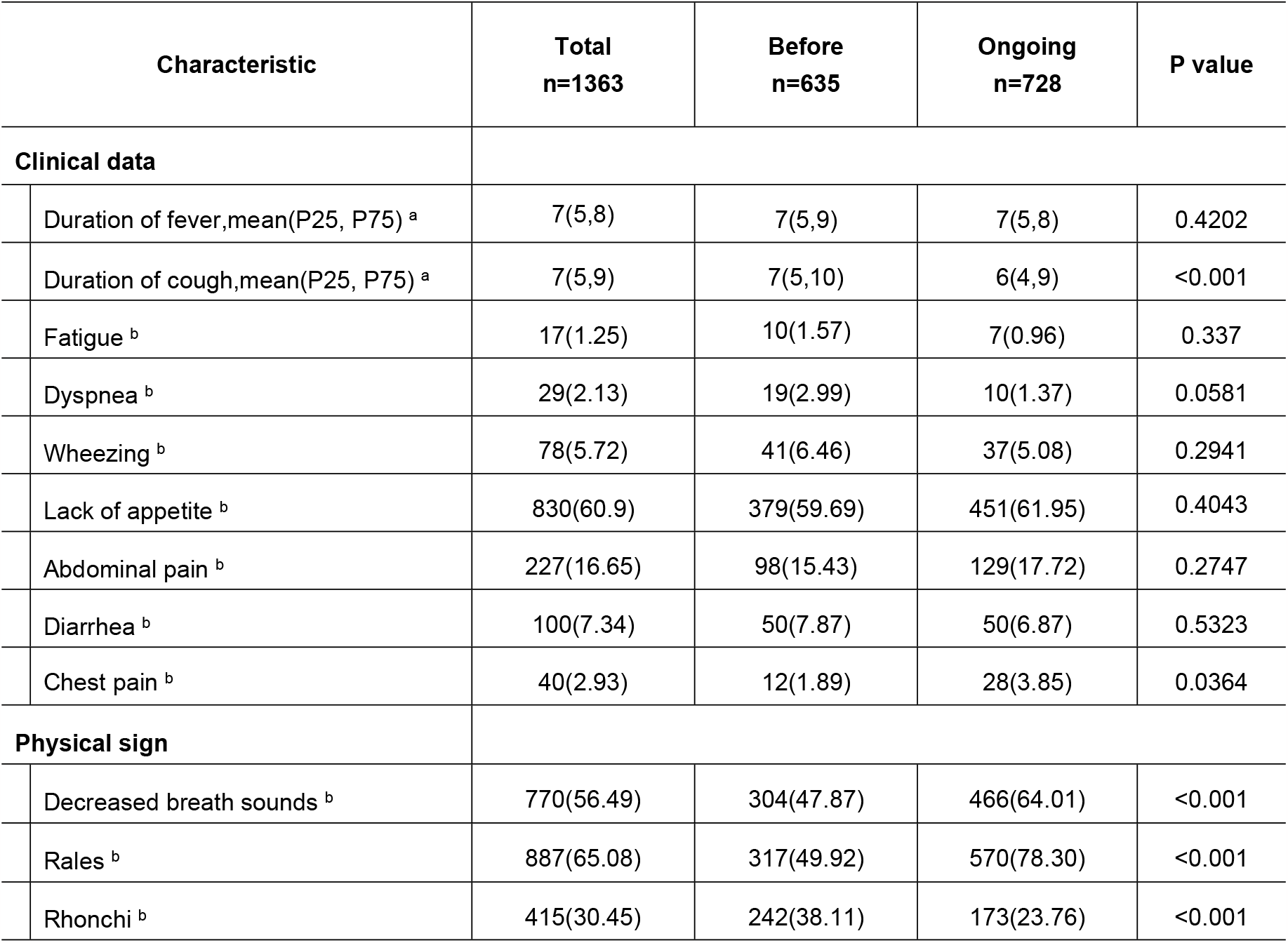
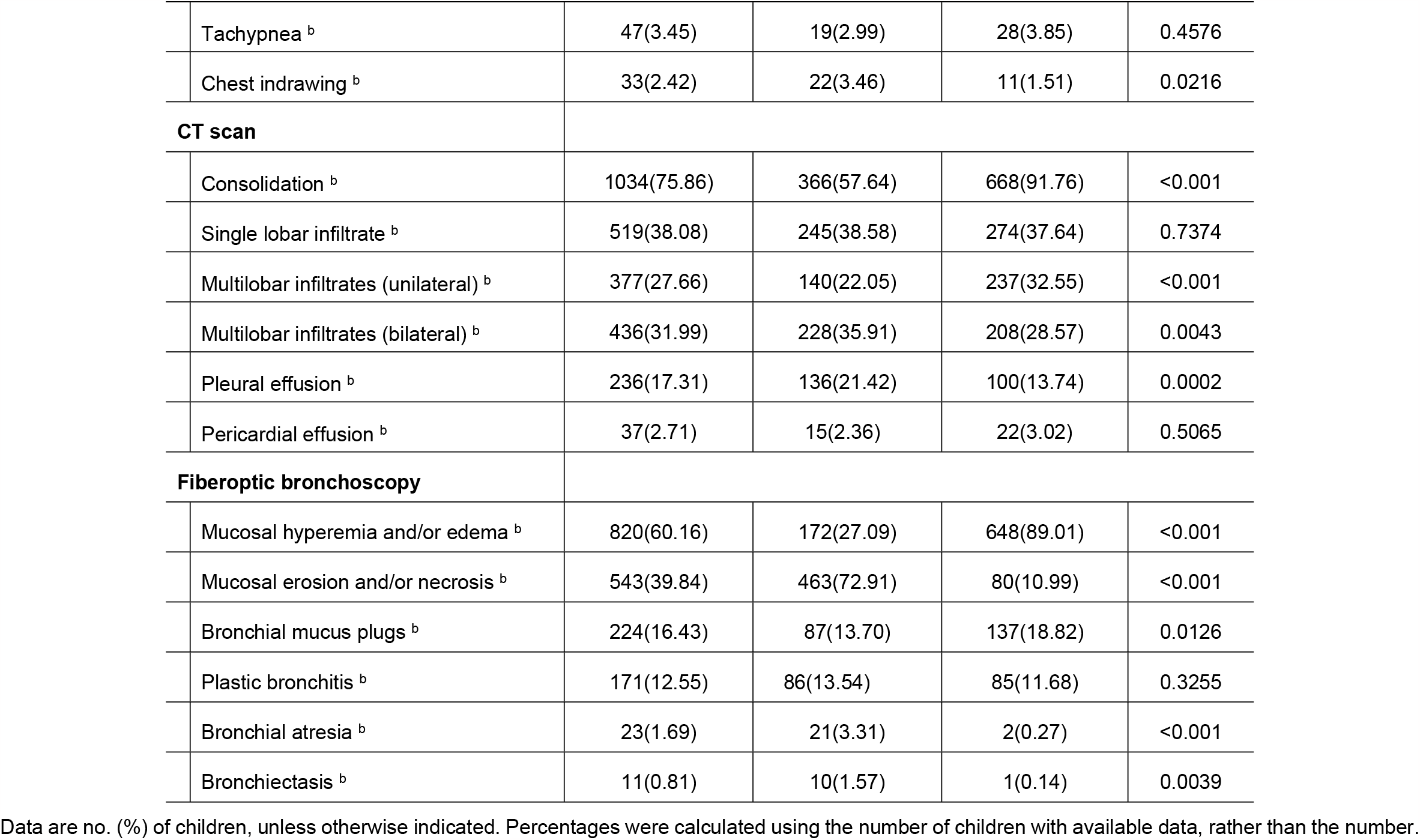
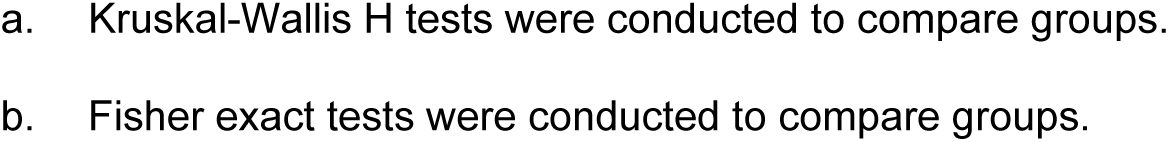
Comparison of clinical and laboratory findings before and during COVID-19 pandemic in children performed with FOB combined with BAL.

| Characteristic | Total<br>n=1363 | Before<br>n=635 | Ongoing<br>n=728 | P value |
| --- | --- | --- | --- | --- |
| **Clinical data** |  |  |  |  |
| Duration of fever,mean(P25, P75) <sup>a</sup> | 7(5,8) | 7(5,9) | 7(5,8) | 0.4202 |
| Duration of cough,mean(P25, P75) <sup>a</sup> | 7(5,9) | 7(5,10) | 6(4,9) | <0.001 |
| Fatigue <sup>b</sup> | 17(1.25) | 10(1.57) | 7(0.96) | 0.337 |
| Dyspnea <sup>b</sup> | 29(2.13) | 19(2.99) | 10(1.37) | 0.0581 |
| Wheezing <sup>b</sup> | 78(5.72) | 41(6.46) | 37(5.08) | 0.2941 |
| Lack of appetite <sup>b</sup> | 830(60.9) | 379(59.69) | 451(61.95) | 0.4043 |
| Abdominal pain <sup>b</sup> | 227(16.65) | 98(15.43) | 129(17.72) | 0.2747 |
| Diarrhea <sup>b</sup> | 100(7.34) | 50(7.87) | 50(6.87) | 0.5323 |
| Chest pain <sup>b</sup> | 40(2.93) | 12(1.89) | 28(3.85) | 0.0364 |
| **Physical sign** |  |  |  |  |
| Decreased breath sounds <sup>b</sup> | 770(56.49) | 304(47.87) | 466(64.01) | <0.001 |
| Rales <sup>b</sup> | 887(65.08) | 317(49.92) | 570(78.30) | <0.001 |
| Rhonchi <sup>b</sup> | 415(30.45) | 242(38.11) | 173(23.76) | <0.001 |
| Tachypnea <sup>b</sup> | 47(3.45) | 19(2.99) | 28(3.85) | 0.4576 |
| Chest indrawing <sup>b</sup> | 33(2.42) | 22(3.46) | 11(1.51) | 0.0216 |
| **CT scan** |  |  |  |  |
| Consolidation <sup>b</sup> | 1034(75.86) | 366(57.64) | 668(91.76) | <0.001 |
| Single lobar infiltrate <sup>b</sup> | 519(38.08) | 245(38.58) | 274(37.64) | 0.7374 |
| Multilobar infiltrates (unilateral) <sup>b</sup> | 377(27.66) | 140(22.05) | 237(32.55) | <0.001 |
| Multilobar infiltrates (bilateral) <sup>b</sup> | 436(31.99) | 228(35.91) | 208(28.57) | 0.0043 |
| Pleural effusion <sup>b</sup> | 236(17.31) | 136(21.42) | 100(13.74) | 0.0002 |
| Pericardial effusion <sup>b</sup> | 37(2.71) | 15(2.36) | 22(3.02) | 0.5065 |
| **Fiberoptic bronchoscopy** |  |  |  |  |
| Mucosal hyperemia and/or edema <sup>b</sup> | 820(60.16) | 172(27.09) | 648(89.01) | <0.001 |
| Mucosal erosion and/or necrosis <sup>b</sup> | 543(39.84) | 463(72.91) | 80(10.99) | <0.001 |
| Bronchial mucus plugs <sup>b</sup> | 224(16.43) | 87(13.70) | 137(18.82) | 0.0126 |
| Plastic bronchitis <sup>b</sup> | 171(12.55) | 86(13.54) | 85(11.68) | 0.3255 |
| Bronchial atresia <sup>b</sup> | 23(1.69) | 21(3.31) | 2(0.27) | <0.001 |
| Bronchiectasis <sup>b</sup> | 11(0.81) | 10(1.57) | 1(0.14) | 0.0039 |

Data are no. (%) of children, unless otherwise indicated. Percentages were calculated using the number of children with available data, rather than the number.
a. Kruskal-Wallis H tests were conducted to compare groups. b. Fisher exact tests were conducted to compare groups.

## Discussion

Respiratory pathogen detection in nasopharyngeal swabs, oropharyngeal swabs and even endotracheal aspirates may not fully represent the etiology of pneumonia[20]. Thus, most children with CAP may not receive treatment based on a reliable etiological diagnosis. Furthermore, the pathogen spectrum of respiratory tract infections is affected by age, geography, and environmental, social, economic and other factors. It is therefore necessary to definitively address the spectrum of respiratory pathogens involved in pediatric CAP. The aim of this study was to evaluate the etiologies in children with CAP requiring BAL. In addition, the influence of the COVID-19 pandemic on the pathogens and the clinical manifestations in the children were analyzed.

We found that the positive rate of pathogen detection was 91.66%. The high etiological detection rate may be mainly attributable to our use of mRT-PCR on specimen from the foci of inflammation. The most common pathogen was *M. pneumoniae*, which was detected in more than four-fifths of all cases, with a remarkable over-representation among children older than 3 years of age. This proportion was higher than that reported in other studies (Lee *et al*. 45.3% in South Korea[4] and Gao *et al*. 37.5% in north China[3]. Considering that patients who required BAL in addition to intravenous medical treatment had more severe pneumonia, this difference may partly be explained by the differences in the studied populations.

Virus was detected in 12.51% (186/1487) of our enrolled cases. Unlike our results, one or more viruses were detected in 73% of children hospitalized for CAP in the United States[21]. Furthermore, a higher percentage of children (94.3%) were reported with severe acute respiratory infection in China during the 2009 H1N1 pandemic and post-pandemic period[22]. With the differences in study sites, populations, as well as study years and duration, the proportions varied largely between studies. Of the 186 children infected with virus, AdV was the most prevalent in the present study representing 34.41% (64/186) of cases, which was in line with a study in Cameroon among children with severe acute respiratory infections[23]. However, the majority of similar studies reported RSV as the most commonly detected viral pathogen[20,21]. In the present study, RSV ranked fifth with a relatively low positive detection rate of 10.22% (19/186). Co-infections were observed in nearly half of the patients (42.47%, 79/186) that tested positive for virus infection. This suggests that severe viral-related CAP is often accompanied by atypical bacterial infection.

The COVID-19 pandemic and the NPIs undertaken during the pandemic significantly affected the transmission of common respiratory pathogens. Compared with before the COVID-19 pandemic, the overall detection rate of viral pathogens was lower during the COVID-19 pandemic. However, the detection rate of atypical bacterial pathogens increased. The decreased detection of AdV, HRV, Flu B, Flu A and HBoV greatly contributed to the overall reduction in viral pathogens. In accordance with our results, a significant decrease in the detection of AdV, Flu B and Flu A was also observed in children with a lower respiratory tract infection during the COVID-19 pandemic in southern China[5] and a considerable reduction in all viral respiratory infections was observed in a large Italian tertiary hospital[6]. In a study conducted in southern China, the detection rates of *M. pneumoniae* using nasopharyngeal swabs of inpatients with acute respiratory infection were also unprecedentedly reduced from 20% in 2019 to 1.0% in 2020[7]. However, another study using BALF from a city in northern China, showed that *M. pneumoniae* was the most frequently detected pathogen, at a rate of up to 73.55%[24]. Furthermore, the detection of *M. pneumoniae* further increased despite the NPIs that were adopted during the COVID-19 pandemic. The main reason for this phenomenon might be the long-term colonization of *M. pneumoniae* in the upper respiratory tract[25].

In aspect to clinical manifestations, we found that children with atypical bacterial infections more frequently displayed a longer duration of fever, and a higher proportion of decreased breath sounds and rales. However, dyspnea, wheezing, diarrhea, rhonchi and chest indrawing were more frequent among children with a viral infection. Similar to other studies[26], compared with children with viral pneumonia, *M. pneumoniae-*positive children were significantly more likely to report a long duration of symptoms before hospitalization, and decreased breath sounds and rales, but were less likely to report dyspnea, wheezing, diarrhea, rhonchi and chest indrawing. Such findings may help inform clinicians of the preliminary differentiation between viral and atypical bacterial infections.

On CT scans, consolidation and unilateral lung infiltration were often seen in the atypical bacterial infection group, whereas bilateral lung infiltration was more common in the viral infection group, suggesting that pulmonary inflammation caused by *M. pneumoniae* is relatively limited to unilateral lung tissue, while viral infection affected bilateral lung tissues in nearly half of the children. This finding was consistent with another study that compared the radiographic findings of children hospitalized for CAP with and without *M. pneumoniae* using data from the EPIC study[27]. FB results indicated more serious inflammatory changes in the mucosa, increasing the likelihood of this condition developing into bronchial mucus plugs and even bronchial dendritic casts in *M. pneumoniae*-infected children. The presence of bronchial mucus plugs, as detected by FB, is independently associated with a longer time period to radiographic clearance in patients with refractory *M. pneumoniae* pneumonia[28]. Wang *et al*. reported that in 86.6% of children with plastic bronchitis the causative agent was *M. pneumoniae*[29] and this result was consistent with our findings. Bronchial atresia and/or bronchiectasis of the distal airway mostly occurred in children infected with virus. Despite few articles related to bronchial lumen morphological changes during the acute stage of pneumonia, our experienced bronchoscopists found that bronchial lumen dilation can be reversed with appropriate treatment. It was unfortunate that bronchial atresia was usually not reversible. Thus, the results highlighted the necessity for careful management and long-term follow-up of patients with mucus plugs, plastic bronchitis and bronchial lumen morphological changes.

Several studies have reported the impact of the COVID-19 pandemic on the pathogen spectrum of respiratory tract infections in children[5-7,24], but the corresponding differences in clinical manifestations have rarely been studied. During the COVID-19 pandemic, we found a decreased proportion of rhonchi, chest indrawing, bilateral lung infiltrates and bronchial lumen morphological changes, but a significantly higher proportion of decreased breath sounds, rales, consolidation, unilateral lung infiltrates and bronchial mucus plugs. Considering that the clinical manifestations of the children were consistent with a higher detection rate of *M. pneumoniae*, a possible explanation for this difference was a decrease in viral infection accompanied by an increase in *M. pneumoniae* infection. Therefore, wearing masks and social distancing in public places could effectively reduce the transmission of respiratory viruses, but did not reduce the infection rate of *M. pneumoniae*. Compared with before the COVID-19 pandemic, fewer children with bilateral multilobar infiltrates and pleural effusion were detected by CT scans, and mild inflammatory changes in the mucosa and decreased lumen morphological changes were detected by FB. Therefore, it is reasonable to deduce that the NPIs during the COVID-19 pandemic significantly alleviated lung inflammation compared with before the pandemic.

Several limitations should be taken into consideration in this present study. First, our study was limited to children requiring BALF and did not include children receiving medical therapy alone or in need of intensive care, and therefore does not fully represent the distribution of characteristics of all CAP pathogens. Second, although testing for common respiratory viruses and atypical bacteria, bacterial detection was restricted to the panel of organisms covered by the SureX 13 respiratory pathogen multiplex kit. This may have resulted in underestimation of the overall detection rate for pathogens. Third, due to the implementation of NPIs, our sample collection time was interrupted and were not able to identify the season-dependent changes in the occurrence of CAP caused by each etiology.

Despite these limitations, our study suggest that *M. pneumoniae* infection might be the greatest pediatric disease burden leading to CAP in northern China. Wearing masks and social distancing in public places can effectively reduce the transmission of respiratory viruses, but cannot reduce the infection rate of *M. pneumoniae*. In addition, the NPIs during the COVID-19 pandemic significantly reduced lung inflammation compared with before the pandemic. Our identifying changes in the pathogen spectrum and clinical characteristics might improve clinical management practices and emphasize the protective value of wearing masks and social distancing in children with Community-acquired pneumonia in children.

## Data Availability

All relevant data are within the manuscript.

## Acknowledgments

The authors would like to thank the clinical staff, specifically the medical doctors and nurses, for their help with collecting clinical specimens, and all of the children and their parents for their participation and cooperation.

## References

1. Perin J, Mulick A, Yeung D, Villavicencio F, Lopez G, Strong KL, et al. Global, regional, and national causes of under-5 mortality in 2000-19: An updated systematic analysis with implications for the Sustainable Development Goals. Lancet Child Adolesc Health. 2022;6:106–15.

2. Darby JB, Singh A, Quinonez R. Management of complicated pneumonia in childhood: A review of recent literature. Rev Recent Clin Trials. 2017;12:253–9.

3. Gao LW, Yin J, Hu YH, Liu XY, Feng XL, He JX, et al. The epidemiology of paediatric Mycoplasma pneumoniae pneumonia in North China: 2006 to 2016. Epidemiol Infect. 2019;147:e192.

4. Lee E, Kim CH, Lee YJ, Kim HB, Kim BS, Kim HY, et al. Annual and seasonal patterns in etiologies of pediatric community-acquired pneumonia due to respiratory viruses and Mycoplasma pneumoniae requiring hospitalization in South Korea. Bmc Infect Dis. 2020;20:132.

5. Liu P, Xu M, Cao L, Su L, Lu L, Dong N, et al. Impact of COVID-19 pandemic on the prevalence of respiratory viruses in children with lower respiratory tract infections in China. Virol J. 2021;18:159.

6. Vittucci AC, Piccioni L, Coltella L, Ciarlitto C, Antilici L, Bozzola E, et al. The disappearance of respiratory viruses in children during the COVID-19 pandemic. Int J Environ Res Public Health. 2021;18:9550.

7. Li L, Wang H, Liu A, Wang R, Zhi T, Zheng Y, et al. Comparison of 11 respiratory pathogens among hospitalized children before and during the COVID-19 epidemic in Shenzhen, China. Virol J. 2021;18:202.

8. Takashita E, Kawakami C, Momoki T, Saikusa M, Shimizu K, Ozawa H, et al. Increased risk of rhinovirus infection in children during the coronavirus disease-19 pandemic. Influenza Other Respir Viruses. 2021;15:488–94.

9. Agca H, Akalin H, Saglik I, Hacimustafaoglu M, Celebi S, Ener B. Changing epidemiology of influenza and other respiratory viruses in the first year of COVID-19 pandemic. J Infect Public Health. 2021;14:1186–90.

10. Wan WY, Thoon KC, Loo LH, Chan KS, Oon L, Ramasamy A, et al. Trends in respiratory virus infections during the COVID-19 pandemic in singapore, 2020. JAMA Netw Open. 2021;4:e2115973.

11. Kuitunen I, Artama M, Mäkelä L, Backman K, Heiskanen-Kosma T, Renko M. Effect of social distancing due to the COVID-19 pandemic on the incidence of viral respiratory tract infections in children in finland during early 2020. Pediatr Infect Dis J. 2020;39:e423–7.

12. Collaborators GLRI. Estimates of the global, regional, and national morbidity, mortality, and aetiologies of lower respiratory infections in 195 countries, 1990-2016: A systematic analysis for the Global Burden of Disease Study 2016. Lancet Infect Dis. 2018;18:1191–210.

13. Yun KW, Wallihan R, Juergensen A, Mejias A, Ramilo O. Community-Acquired pneumonia in children: Myths and facts. Am J Perinatol. 2019;36:S54–7.

14. Radhakrishnan D, Yamashita C, Gillio-Meina C, Fraser DD. Translational research in pediatrics III: Bronchoalveolar lavage. Pediatrics. 2014;134:135–54.

15. Wang L, Zhao M, Shi Z, Feng Z, Guo W, Yang S, et al. A GeXP-Based assay for simultaneous detection of multiple viruses in hospitalized children with community acquired pneumonia. Plos One. 2016;11:e162411.

16. Midulla F, de Blic J, Barbato A, Bush A, Eber E, Kotecha S, et al. Flexible endoscopy of paediatric airways. Eur Respir J. 2003;22:698–708.

17. Experts Group of Pediatric Respiratory Endoscopy, Talent Exchange Serice Center of National Health Commission Endoscopy Committee, Pediatric Section of Chinese Medical Doctor Association, Pediatric Respiratory Endoscopy Committee, Endoscopists Section of Chinese Medical Doctor Association, Pediaric Interventional Respirology Group, Maternal and Pediatric Minimally Inasive Section of Chinese Maternal and Child Health Association Bronchoscopy Collaboration Subgroup of Respirology Group, Pediatric Section of Chinese Medical Associatio. Guideline of pediatric flexible bronchoscopy in China (2018 version). Zhong Hua Shi Yong Er Ke Lin Chuang Za Zhi. 2018;13:983–989.

18. Chen D, Huang Y, Jiao A, Jin R, Liu X, Meng C, et al. Expert consensus on respiratory endoscopic interventional diagnosis and treatment of refractory pneumonia in Chinese children. Zhong Hua Shi Yong Er Ke Za Zhi. 2019;34:449–57.

19. Andronikou S, Goussard P, Sorantin E. Computed tomography in children with community-acquired pneumonia. Pediatr Radiol. 2017;47:1431–40.

20. Rhedin S, Lindstrand A, Hjelmgren A, Ryd-Rinder M, Ohrmalm L, Tolfvenstam T, et al. Respiratory viruses associated with community-acquired pneumonia in children: Matched case-control study. Thorax. 2015;70:847–53.

21. Jain S, Williams DJ, Arnold SR, Ampofo K, Bramley AM, Reed C, et al. Community-acquired pneumonia requiring hospitalization among U.S. Children. N Engl J Med. 2015;372:835–45.

22. Zhang C, Zhu N, Xie Z, Lu R, He B, Liu C, et al. Viral etiology and clinical profiles of children with severe acute respiratory infections in China. Plos One. 2013;8:e72606.

23. Kenmoe S, Tchendjou P, Vernet MA, Moyo-Tetang S, Mossus T, Njankouo-Ripa M, et al. Viral etiology of severe acute respiratory infections in hospitalized children in Cameroon, 2011-2013. Influenza Other Respir Viruses. 2016;10:386–93.

24. Guo W, Cui X, Wang Q, Wei Y, Guo Y, Zhang T, et al. Clinical evaluation of metagenomic next-generation sequencing for detecting pathogens in bronchoalveolar lavage fluid collected from children with community-acquired pneumonia. Front Med (Lausanne). 2022;9:952636.

25. Spuesens EB, Fraaij PL, Visser EG, Hoogenboezem T, Hop WC, van Adrichem LN, et al. Carriage of Mycoplasma pneumoniae in the upper respiratory tract of symptomatic and asymptomatic children: An observational study. Plos Med. 2013;10:e1001444.

26. Li YT, Liang Y, Ling YS, Duan MQ, Pan L, Chen ZG. The spectrum of viral pathogens in children with severe acute lower respiratory tract infection: A 3-year prospective study in the pediatric intensive care unit. J Med Virol. 2019;91:1633–42.

27. Kutty PK, Jain S, Taylor TH, Bramley AM, Diaz MH, Ampofo K, et al. Mycoplasma pneumoniae Among Children Hospitalized with Community-acquired Pneumonia. Clin Infect Dis. 2019;68:5–12.

28. Huang L, Huang X, Jiang W, Zhang R, Yan Y, Huang L. Independent predictors for longer radiographic resolution in patients with refractory Mycoplasma pneumoniae pneumonia: A prospective cohort study. Bmj Open. 2018;8:e23719.

29. Wang L, Wang W, Sun JM, Ni SW, Ding JL, Zhu YL, et al. Efficacy of fiberoptic bronchoscopy and bronchoalveolar lavage in childhood-onset, complicated plastic bronchitis. Pediatr Pulmonol. 2020;55:3088–95.

